# Prevalence of SARS-CoV-2 Antibodies after the Omicron Surge, Kingston, Jamaica, 2022

**DOI:** 10.1101/2022.09.20.22280173

**Authors:** Joshua J. Anzinger, Suzette M. Cameron-McDermott, Yakima Z.R. Phillips, Leshawn Mendoza, Mark Anderson, Gavin Cloherty, Susan Strachan-Johnson, John F. Lindo, J. Peter Figueroa

## Abstract

A cross-sectional SARS-CoV-2 serosurvey was conducted after the Omicron surge in Jamaica using 1,540 samples collected during March – May 2022 from persons attending antenatal, STI and non-communicable diseases clinics in Kingston, Jamaica. SARS-CoV-2 spike receptor binding domain (RBD) and/or nucleocapsid IgG antibodies were detected for 88.4% of the study population, with 77.0% showing evidence of previous SARS-CoV-2 infection. Of persons previously infected with SARS-CoV-2 and/or with COVID-19 vaccination, 9.6% were negative for spike RBD IgG, most of which were unvaccinated previously infected persons. Amongst unvaccinated previously infected people, age was associated with testing spike RBD IgG negative. When considering all samples, median spike RBD IgG levels were 131.6 BAU/mL for unvaccinated persons with serological evidence of past infection, 90.3 BAU/mL for vaccinated persons without serological evidence of past infection, and 896.1 BAU/mL for vaccinated persons with serological evidence of past infection. Our study of the first reported SARS-CoV-2 serosurvey in Jamaica shows extensive SARS-CoV-2 population immunity, identifies a substantial portion of the population lacking spike RBD IgG, and provides additional evidence for increasing COVID-19 vaccine coverage in Jamaica.

## 1. Background

Jamaica has the second lowest COVID-19 vaccine coverage (26.7% fully vaccinated as of September 6, 2022) in the Western Hemisphere [1] and relies on COVID-19 case reporting to inform the extent of SARS-CoV-2 infections in the population. Although virological testing (i.e., PCR and rapid antigen) increased throughout the pandemic in Jamaica, even in highly resourced countries most cases were not identified [2,3]. Serosurveys provide a more accurate estimation of the extent of SARS-CoV-2 infections than virological testing and can aid in guiding public health decisions.

Antibodies against SARS-CoV-2 are useful to determine past infection and to potentially identify persons at risk of COVID-19. In the context of a population with spike-encoding COVID-19 vaccines such as Jamaica, SARS-CoV-2 nucleocapsid IgG can be used to identify persons infected with SARS-CoV-2 in the past [4]. Spike IgG antibodies are the most clinically important type of SARS-CoV-2 antibodies as they can prevent SARS-CoV-2 from binding to ACE-2 receptors and in the context of Omicron have been shown to be associated with a more favorable clinical course of COVID-19 [5].

After the initial surge of COVID-19 during September 2020, Jamaica experienced an Alpha surge during February – April 2021, a Delta surge during August – October 2021, and then an Omicron surge during December 2021 – February 2022 [1,6]. Despite these multiple surges over two years, the extent of population SARS-CoV-2 immunity is unclear as there have been no reported serosurveys of the Jamaican population.

## 2. Objectives

To determine the level of persons previously infected with SARS-CoV-2 and the extent of evidence of humoral immunity using assays for SARS-CoV-2 nucleocapsid and spike IgG antibodies in the population of Kingston, Jamaica.

## 3. Study Design

### 3.1 Study Participants

We determined the presence of SARS-CoV-2 nucleocapsid and spike IgG in residual serum samples submitted during March 16 – May 5, 2022, from the Comprehensive Health Centre (CHC) Antenatal (ANC), Non-communicable Diseases (NCD) and Sexually Transmitted Infections (STI) Clinics. Samples for this cross-sectional study were collected and tested irrespective of age except for the NCD clinic in which only samples from persons ≥50 years of age were selected to increase the number of participants in this study of that age group. The clinics were targeted to represent the adult population of the Kingston Metropolitan Area by including both sexes, pregnant women, and a broad range of ages. NCD and STI clinic participants were 25.5% and 39.3% male, respectively. Participants provided self-reported previous COVID-19 and COVID-19 vaccination history. Booster information and COVID-19 vaccine type received was not available. As of September 6, 2022, in Jamaica 727,406 people were fully vaccinated, 828,809 received one dose, and 44,234 received a booster dose. The percentages of COVID-19 vaccine doses administered in Jamaica as of September 6, 2022 were: AstraZeneca (ChAdOx1), 56%; Pfizer-BioNTech (BNT162b2), 36%; Johnson and Johnson (Ad26.COV2.S), 8%; and Sinopharm (BBIBP-CorV), 0.06%. No other COVID-19 vaccine types were administered in the public sector.

### 3.2 Procedures

A total of 1540 unique patient samples (335 for ANC, 374 for STI, and 831 for NCD) were tested using chemiluminescence immunoassay (CMIA) using the Abbott ARCHITECT SARS-CoV-2 IgG assay for nucleocapsid-specific IgG and the AdviseDx SARS-CoV-2 IgG II Quant assay for spike-specific receptor binding domain (RBD) IgG. All tests were performed using an ARCHITECT *i*1000SR instrument. The manufacturer’s recommended cutoff of ≥50.0 arbitrary units/milliliter (AU/mL) was used for the SARS-CoV-2 IgG II Quant assay and a lower cutoff of ≥0.4 signal-to-cutoff (S/CO) was used for the SARS-CoV-2 IgG assay to maximize sensitivity [7,8]. To convert spike RBD IgG AU/mL to international binding antibody units/milliliter (BAU/mL), spike IgG AU/mL units were multiplied by 0.142. This study was approved by the Ministry of Health and Wellness Ethics Committee (2022/20).

### 3.3 Statistical Analysis

Comparison of antibody levels among groups was done with a one-way ANOVA with Tukey post hoc test for multiple comparisons using GraphPad Prism version 9.4.1 (GraphPad Software, San Diego, California, USA). Categorical variables were assessed using the χ^2^ test and correlations were determined by Pearson’s correlation, both analyzed using IBM SPSS Statistics for Windows version 20 (IBM Corp., Armonk, New York, USA).

## 4. Results

Participants were assessed for previous COVID-19, COVID-19 vaccination, and SARS-CoV-2 IgG antibodies (Table 1). Only 4.7% of all participants reported previous COVID-19 with similar percentages between clinics. COVID-19 vaccination coverage was significantly different between clinics (χ^2^ = 100.12, *p* < 0.0001), with 16.7% for ANC, 45.4% for NCD, and 26.7% for STI, coinciding with a significantly different (χ^2^ = 1543.77, *p* < 0.0001) mean age between ANC (26.9 ± 6.2), NCD (65.1 ± 9.7), and STI (33.6 ± 14.9) clinic attendees. The overall prevalence of nucleocapsid IgG and spike RBD IgG was 69.1% and 81.4%, respectively, with 88.4% positive for either nucleocapsid IgG or spike RBD IgG. Spike RBD IgG and nucleocapsid IgG were correlated for each of the clinics (Supplemental Figure). For each clinic the prevalence of nucleocapsid and spike RBD IgG antibodies, respectively, was 70.4% and 81.5% for ANC, 65.2% and 79.8% for NCD, and 76.5% and 85.0% for STI. Age showed a weak inverse association (*r* = -0.118, <0.0001) with testing nucleocapsid IgG positive, and females were more likely than males to be nucleocapsid IgG positive (χ^2^ = 9.827, *p* = 0.002) but were similar to males for testing spike RBD IgG positive (χ^2^ = 3.083, *p* = 0.079). Neither age nor sex were associated with being unvaccinated without previous infection when compared to unvaccinated persons with previous infection and all vaccinated persons as a single group. We also assessed the percentage of unvaccinated persons with spike RBD IgG but without nucleocapsid IgG to more accurately reflect the total number of people with serological evidence of previous infection. For ANC, NCD, and STI clinics the percentage of unvaccinated persons with spike RBD IgG but without nucleocapsid IgG was 16.1%, 4.7%, and 7.5%, respectively. Combining these percentages with the percentage of persons testing nucleocapsid IgG positive resulted in 86.5% for ANC, 69.9% for NCD, and 84.0% for STI having serological evidence of having previously been infected with SARS-CoV-2, with 77.0% overall when all sub-populations were considered together.

**Table 1.** Prevalence of self-reported COVID-19, COVID-19 vaccination, and SARS-CoV-2 antibodies

|  |  |  |  |  |  |  |  |  |
| --- | --- | --- | --- | --- | --- | --- | --- | --- |
| Clinic, variable | Observations, no. | COVID-19, no. (%) | Vaccination, no. (%) | S IgG Positive, no. (%) | NC IgG Positive, no. (%) | S or NC IgG Positive, no. (%) | Unvaccinated, NC IgG Negative, S IgG Positive, no (%) | Evidence of Previous COVID-19, no. (%) |
| **ANC** |  |  |  |  |  |  |  |  |
| Age, y |  |  |  |  |  |  |  |  |
| <20 | 35 | 3* (8.6%) | 5 (14.3%) | 27 (77.1%) | 25 (71.4%) | 30 (85.7%) | 3 (8.5%) | 28 (80.0%) |
| 20-29 | 193 | 5 (2.6%) | 23 (11.9%) | 158 (81.9%) | 137 (71.0%) | 175 (90.7%) | 34 (17.6%) | 171 (88.6%) |
| 30-39 | 98 | 7 (7.1%) | 25 (25.5%) | 80 (81.6%) | 69 (70.4%) | 88 (89.8%) | 15 (15.3%) | 84 (85.7%) |
| ≥40 | 9 | 0 (0.0%) | 3 (33.3%) | 8 (88.9%) | 5 (55.6%) | 8 (88.9%) | 2 (22.2%) | 7 (77.8%) |
| Total | 335 | 15 (4.5%) | 56 (16.7%) | 273 (81.5%) | 236 (70.4%) | 301 (90.0%) | 54 (16.1%) | 290 (86.5%) |
| **NCD** |  |  |  |  |  |  |  |  |
| Age, y |  |  |  |  |  |  |  |  |
| 50-59 | 286 | 10 (3.5%) | 125 (43.7%) | 233 (81.5%) | 199 (69.6%) | 249 (87.1%) | 12 (4.2%) | 211 (73.8%) |
| 60-69 | 293 | 18 (6.1%) | 135 (46.1%) | 240 (81.9%) | 189 (64.5%) | 260 (88.7%) | 18 (6.1%) | 207 (70.6%) |
| 70-79 | 180 | 7 (3.8%) | 85 (47.2%) | 144 (80.0%) | 113 (62.8%) | 156 (86.7%) | 7 (3.9%) | 120 (66.7%) |
| ≥80 | 72 | 3* (4.2%) | 32 (44.4%) | 46 (63.9%) | 41 (56.9%) | 55 (76.4%) | 2 (2.8%) | 43 (59.7%) |
| Total | 831 | 38 (4.6%) | 377 (45.4%) | 663 (79.8%) | 542 (65.2%) | 720 (86.6%) | 39 (4.7%) | 581 (69.9%) |
| Sex |  |  |  |  |  |  |  |  |
| M | 212 | 6 (2.8%) | 98 (46.2%) | 164 (77.4%) | 118 (55.7%) | 178 (84.0%) | 13 (6.1%) | 131 (61.8%) |
| F | 619 | 32* (5.2%) | 279 (45.1%) | 499 (80.6%) | 424 (68.5%) | 542 (87.6%) | 26 (4.2%) | 450 (72.7%) |
| **STI** |  |  |  |  |  |  |  |  |
| Age, y |  |  |  |  |  |  |  |  |
| <20 | 60 | 0 (0.0%) | 15 (25.0%) | 54 (90.0%) | 52 (86.7%) | 57 (95.0%) | 0 (0.0%) | 52 (86.7%) |
| 20-29 | 135 | 9@ (6.7%) | 32 (23.7%) | 118 (87.4%) | 104 (77.0%) | 126 (93.3%) | 14 (10.4%) | 118 (87.4%) |
| 30-39 | 69 | 6 (8.7%) | 20 (29.0%) | 61 (88.4%) | 58 (84.1%) | 65 (94.2%) | 4 (5.8%) | 62 (89.9%) |
| 40-49 | 49 | 2 (4.1%) | 8 (16.3%) | 34 (69.4%) | 32 (65.3%) | 38 (77.6%) | 5 (10.2%) | 37 (75.5%) |
| 50-59 | 33 | 1 (3.0%) | 14 (42.4%) | 26 (78.8%) | 20 (60.6%) | 29 (87.8%) | 4 (12.1%) | 24 (72.7%) |
| 60-69 | 21 | 2 (9.5%) | 9 (42.9%) | 19 (90.5%) | 15 (71.4%) | 20 (95.2%) | 1 (4.8%) | 16 (76.2%) |
| 70-79 | 5 | 0 (0.0%) | 2 (40.0%) | 5 (100.0%) | 4 (80.0%) | 5 (100.0%) | 0 (0.0%) | 4 (80.0%) |
| ≥80 | 2 | 0 (0.0%) | 0 (0.0%) | 1 (50.0%) | 1 (50.0%) | 1 (50.0%) | 0 (0.0%) | 1 (50.0%) |
| Total | 374 | 20 (5.3%) | 100 (26.7%) | 318 (85.0%) | 286 (76.5%) | 341 (91.2%) | 28 (7.5%) | 314 (84.0%) |
| Sex |  |  |  |  |  |  |  |  |
| M | 147 | 9* (6.1%) | 40 (27.2%) | 117 (79.6%) | 106 (72.1%) | 134 (91.2%) | 11 (7.5%) | 117 (79.6%) |
| F | 227 | 11* (4.8%) | 60 (26.4%) | 201 (88.5%) | 180 (79.3%) | 207 (91.2%) | 17 (7.5%) | 197 (86.8%) |
| Overall | 1540 | 73 (4.7%) | 533 (34.6%) | 1254 (81.4%) | 1064 (69.1%) | 1362 (88.4%) | 121 (7.9%) | 1185 (77.0%) |

NC = nucleocapsid. S = spike. \*One respondent answered “not sure” and was included as no previous COVID-19. @Two respondents answered “not sure” and were included as no previous COVID-19.

In addition to identifying the percentage of persons without humoral antibody immunity due to COVID-19 vaccination and/or previous SARS-CoV-2 infection (Table 1), we sought to distinguish between persons without detectable spike RBD IgG from those with serological evidence of previous infection (i.e., nucleocapsid IgG positive) and/or COVID-19 vaccination (Table 2). Of the 1,263 persons with previous infection and/or vaccination, 121 (9.6%) were spike RBD IgG negative. Unvaccinated persons with evidence of previous infection represented 88.4% of persons that were spike RBD IgG negative, with 100.0% for ANC, 73.7% for NCD and 92.0% for STI. When comparing persons from all clinics that tested spike RBD IgG positive or negative amongst unvaccinated persons with evidence of previous infection, sex was not whereas age was significantly associated with testing spike RBD IgG negative (χ^2^ = 98.280, *p* = 0.044). Vaccinated persons without evidence of previous infection and vaccinated persons with evidence of previous infection represented 17.4% and 0.8%, respectively, of vaccinated and/or previously infected persons without detectable spike RBD IgG. Only one vaccinated person with evidence of previous infection did not show detectable spike RBD IgG.

**Table 2.** Persons without detectable spike-specific RBD IgG amongst those with COVID-19 vaccination and/or positive for nucleocapsid IgG

| Clinic | Observations,<br>no. | Unvaccinated with<br>previous infection,<br>no. (%) | Vaccinated without<br>previous infection,<br>no. (%) | Vaccinated with<br>previous infection,<br>no. (%) |
| --- | --- | --- | --- | --- |
| ANC | 28 | 28 (100.0%) | 0 (0.0%) | 0 (0.0%) |
| NCD | 76 | 56 (73.7%) | 19 (25%) | 1 (1.3%) |
| STI | 25 | 23 (92.0%) | 2 (8.0%) | 0 (0.0%) |
| Overall | 121 | 107 (88.4%) | 21 (17.4%) | 1 (0.8%) |

We next examined the quantity of spike RBD IgG antibodies for all samples. When stratified by past SARS-CoV-2 infection (i.e., nucleocapsid IgG results) and COVID-19 vaccination (Figure 1), across all clinics, vaccinated persons with serological evidence of past infection showed significantly greater levels of SARS-CoV-2 spike RBD IgG (*p* < 0.0001), compared to unvaccinated persons with serological evidence of past infection, and vaccinated persons without serological evidence of past infection (*p* < 0.0001), except for ANC that was not significantly different (*p* = 0.23). Considering all participants or individual clinics, there was no significant difference in spike RBD IgG levels between unvaccinated persons with serological evidence of past infection and vaccinated persons without serological evidence of past infection. When considering all clinics, median SARS-CoV-2 spike RBD IgG levels for each of the groups were as follows: unvaccinated persons without serological evidence of past infection, 15.6 AU/mL (2.2 bAU/mL); unvaccinated persons with serological evidence of past infection, 926.7 AU/mL (131.6 bAU/mL); vaccinated persons without serological evidence of past infection, 635.8 AU/mL (90.3 bAU/mL); and vaccinated persons with serological evidence of past infection, 6,310.6 AU/mL (896.1 bAU/mL). Stratification of SARS-CoV-2 spike RBD IgG levels by age showed that median spike RBD IgG antibody levels were lower for persons ≥80 years of age compared to other groups, though the difference was not significantly different (Figure 2). However, age showed a weak inverse association (*r* = -0.075, *p* = 0.003) with testing spike RBD IgG positive. When the ≥80 years of age population was compared between vaccination status and previous infection groups (Figure 2), vaccinated persons with serological evidence of past infection showed significantly (*p* <0.01) greater spike RBD IgG antibody levels than unvaccinated previously infected and vaccinated previously uninfected groups.

**Figure 1.**
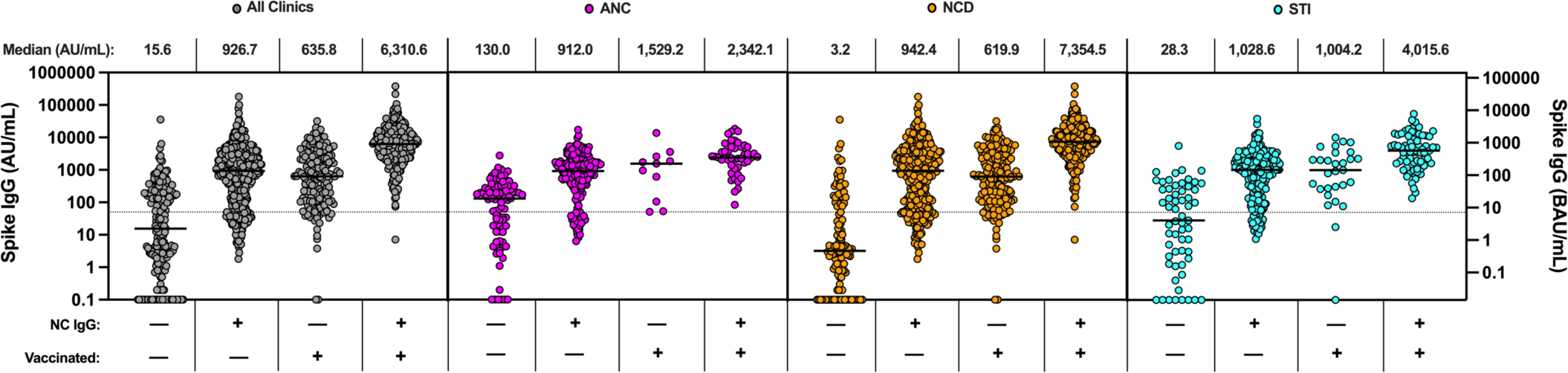
SARS-CoV-2 spike RBD IgG index values of participants stratified for nucleocapsid IgG results and COVID-19 vaccination status for all clinics and individual clinics (ANC, NCD, and STI). Dotted horizontal lines represent the spike RBD IgG test cutoff value of 50 AU/mL. AU/mL = arbitrary units per milliliter. BAU/mL binding antibody units per milliliter.

**Figure 2.**
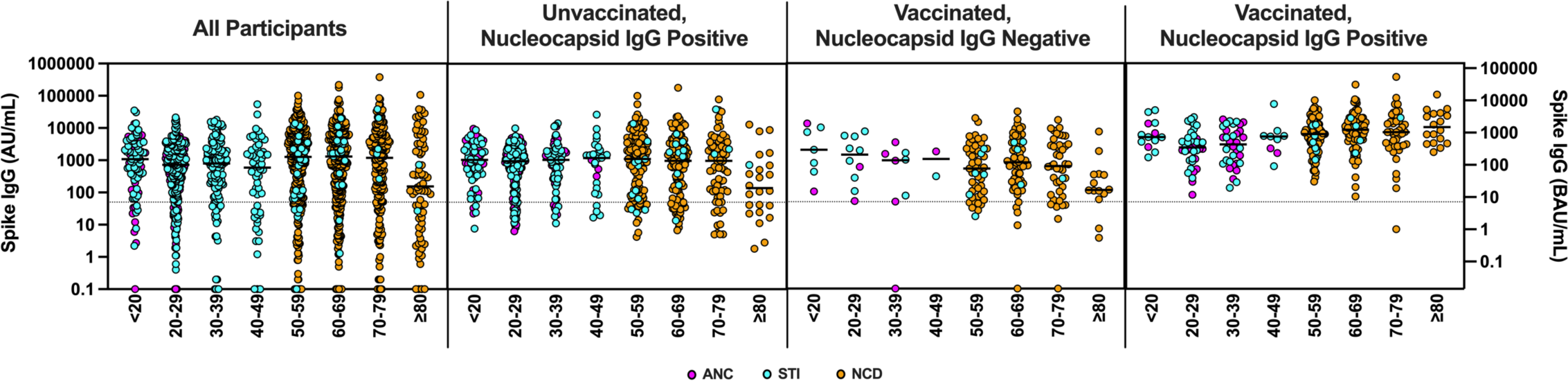
SARS-CoV-2 spike RBD IgG index values of participants stratified by age. Clinics were assessed for all participants, unvaccinated participants testing nucleocapsid IgG positive, vaccinated participants testing nucleocapsid IgG negative, and vaccinated participants testing nucleocapsid IgG positive. Dotted horizontal lines represent the spike RBD IgG test cutoff value of 50 AU/mL. AU/mL = arbitrary units per milliliter. BAU/mL = binding antibody units per milliliter.

## 5. Discussion

Despite low COVID-19 vaccination coverage and low self-reporting of previous COVID-19, our results show that 88% of the Kingston population examined had SARS-CoV-2 IgG antibodies. These findings have important implications for future SARS-CoV-2 circulation and COVID-19 as described below.

Only 4.7% of persons reported previous COVID-19, yet 69% were positive for nucleocapsid IgG and 77% showed serological evidence of previous infection, indicating a considerable level of unrecognized/asymptomatic infections, an important driver of SARS-CoV-2 circulation [9]. Serological evidence of those previously infected with SARS-CoV-2 is most likely an underestimate as seroreversion of nucleocapsid IgG has been reported for the assay used [8,10–12], making it likely that some persons infected in the past tested nucleocapsid IgG negative. A previous study concluded that in previously infected people, starting from the highest level of nucleocapsid IgG detected with the Abbott Architect system (the same test used in our study), nucleocapsid IgG becomes negative 137 days later [13], though this timeframe is likely much greater for our study as we used a lower cutoff value. In addition to nucleocapsid IgG seroreversion, SARS-CoV-2 infected people that failed to seroconvert and those that seroreverted for both nucleocapsid and spike RBD IgG antibodies would also not be identified as previously infected in this study. Regardless of the likely underestimate of previously infected people in our study, our data shows a considerably greater number of people infected compared to virological data, as expected. According to the Jamaican Ministry of Health and Wellness, as of May 5, 2022 (the last day we collected samples for this study) there were 34,735 COVID-19 cases in the Kingston and St. Andrew parishes [14], equating to 6.1% of the population infected (though this number may be slightly inflated due to re-infections), dramatically lower than the 77% of the population we show was previously infected.

In addition to unvaccinated persons without previous SARS-CoV-2 infection that are immunologically naíve and at higher risk of severe disease compared to persons with evidence of immunity, we also identified unvaccinated, previously infected people as the most common group without detectable spike RBD IgG. This likely places them at increased risk for SARS-CoV-2 infection and COVID-19 compared to those with detectable spike RBD IgG [15,16]. These persons were older in age with lower levels of nucleocapsid IgG (data not shown) which may indicate infections in the past but could also be due to a less robust antibody response to infection. These findings are consistent with older age being associated with a less robust immune response to SARS-CoV-2 infection [17] and indicate that a targeted COVID-19 vaccine approach to increase coverage in the elderly population continues to be a priority.

Although levels of SARS-CoV-2 spike IgG have previously been linked to protection from infection and disease (i.e., a correlate of protection), these studies were performed prior to the emergence of Omicron [18]. It has now been widely reported that even for persons with immunity (via infection and/or vaccination) to SARS-CoV-2, breakthrough and/or re-infections are much more likely with Omicron than with previous SARS-CoV-2 variants of concern. A large study in Denmark showed no difference between the rate of breakthrough infections with Omicron comparing the highest and lowest quintiles of spike IgG antibodies [19], whereas a study in South Africa showed that SARS-CoV-2 spike IgG levels of >1549 bAU/mL were associated with reduced risk of Omicron infection (adjusted odds ratio of 0.42) [20]. Regardless of the discrepancy between these studies, only 9.5% of participants in our study had spike IgG levels >1549 bAU/mL, suggesting that the overwhelming majority of persons are likely susceptible to future infections with Omicron or a more immune evasive variant. To our knowledge spike RBD IgG levels have not been assessed in the context of SARS-CoV-2 circulation, but spike antibody levels have been shown to be associated with decreased risk of fever, hypoxia and inflammation when infected with Omicron [5].

It is not clear if previous infection or vaccination provided superior protection from COVID-19 but several recent studies suggest that infection, particularly with a closely related variant, provided greater protection, while previous infection with vaccination, also termed hybrid immunity [21], provided superior protection and is consistent with the population in our study having the greatest spike RBD IgG antibody levels we observed. A study examining a representative population of Qatar showed that previous infection was associated with a 46.1% effectiveness against symptomatic Omicron infection, whereas vaccination with an mRNA vaccine showed no effectiveness against symptomatic Omicron infection (though vaccination time from infection was almost exclusively >6 months) [22]. In the same study, boosting with an mRNA vaccine of previously uninfected or infected person resulted in a 55.2% and 77.3% effectiveness against symptomatic infection, respectively. Recently, protection from Omicron infection was associated with previous infection with pre-Omicron variants of concern and even greater with previous infection with Omicron [23], though this study was in the context of a population with nearly universal COVID-19 vaccine coverage. A recent pre-print study showed that previous infection with pre-Omicron variants was associated with 15.1% effectiveness against symptomatic infection, whereas previous infection with an Omicron variant was associated with a 76.1% effectiveness against symptomatic infection with Omicron BA.4 or BA.5 [24]. Although the extent of Omicron infections compared to previous variants in Jamaica is unclear, the high level of previous infection is likely to reduce the risk of a large COVID-19 surge in the near term (as has been observed up to September 7, 2022). However, identification of unvaccinated persons with or without previous infection who lack spike RBD IgG provided evidence for persons at risk of infection and indicated the need for increased COVID-19 vaccine coverage. Our findings together with that of others showing a more robust spike IgG response when vaccinating previously infected persons (hybrid immunity) provides further evidence for increasing vaccine coverage even in a population in which at least 77% of persons have been previously infected. This may be particularly important in persons ≥80 years of age in which spike RBD IgG levels were significantly greater than vaccinated previously infected and unvaccinated previously infected persons ≥80 years of age. Increasing COVID-19 vaccine coverage with variant-specific COVID-19 vaccines would likely be of even greater benefit than ancestral COVID-19 vaccines in the low vaccine coverage setting described as a recent pre-print modeling study concludes [25].

Our study was limited by having only a very small number of children (34) in the study population (with none <13 year old), the lack of Gold Standard live virus neutralizing antibody testing that requires a BSL3 (not present in Jamaica), dates of infection, and more detailed information related to vaccination history (e.g., type and date of vaccination). It is possible that a very small number of persons with nucleocapsid IgG could be due to vaccination with an inactivated vaccine, although only 959 vaccine doses of Sinopharm had been administered in Jamaica, making it unlikely that this would have any meaningful impact of our reported prevalence of previous infection.

In conclusion, our study provides the first evidence for the extent of SARS-CoV-2 immunity due to vaccination and/or infection in the largest city in Jamaica. Our data can guide public health decisions, allowing for a more accurate estimation of COVID-19 risk in the population, better identification of those most at risk of COVID-19, and provides further support for increasing COVID-19 vaccine coverage.

## Data Availability

All data produced in the present study are available upon reasonable request to the authors

## Acknowledgements

As a Global Infectious Diseases Scholar, Suzette Cameron-McDermott received mentored research training in the development of this manuscript. This training was supported in part by the University at Buffalo Clinical and Translational Science Institute award UL1TR001412 and the Global Infectious Diseases Research Training Program award D43TW010919. The content is solely the responsibility of the authors and does not necessarily represent the official views of the Clinical and Translational Science Institute or the National Institutes of Health.

---

**Supplemental Figure.**
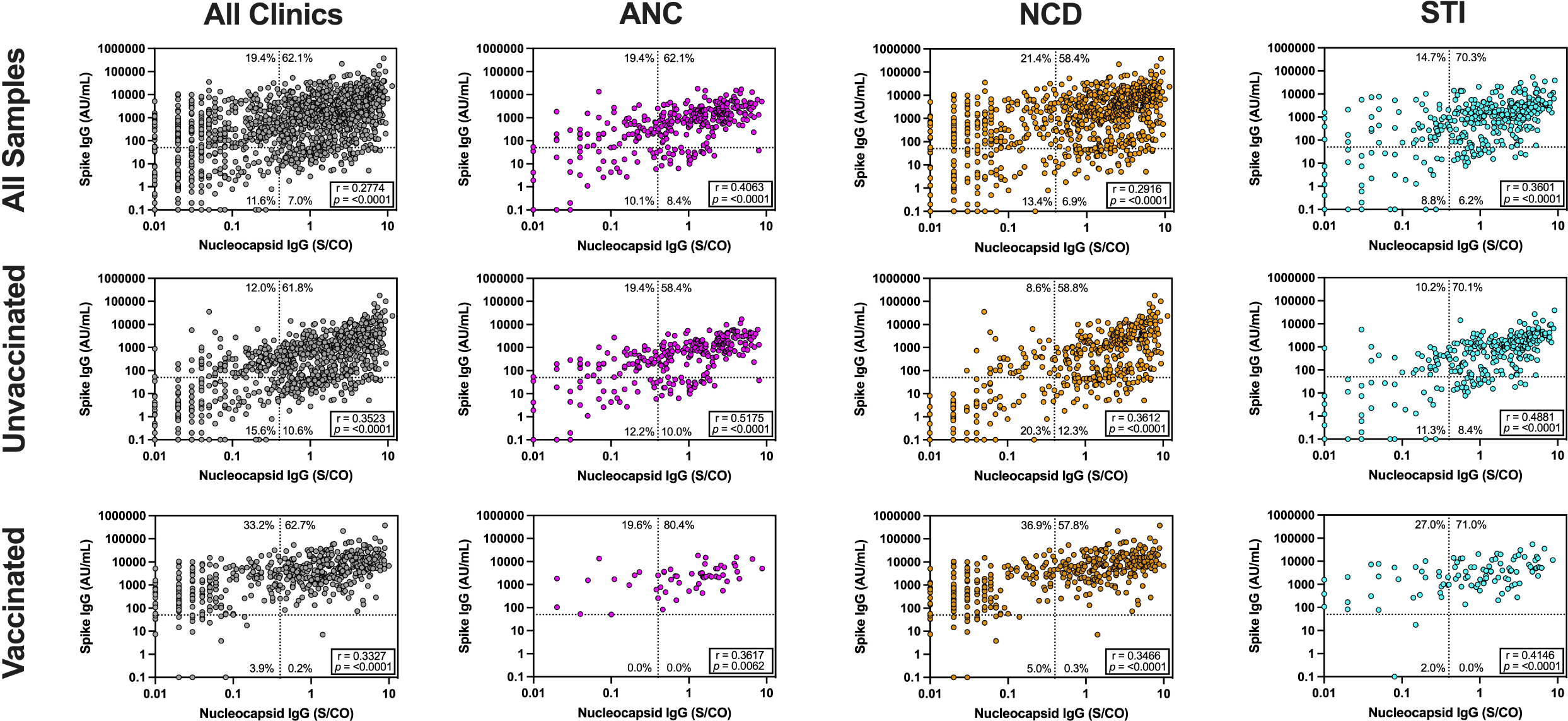
SARS-CoV-2 nuclcocapsid IgG and spike RBD IgG index values of participants for all clinics and individual clinics (ANC, NCD, and STI). Dotted lines represent the test cutoff values (50 AU/mL for spike RBD IgG and 0.4 S/CO for nucleocapsid IgG). AU/mL = arbitrary units per milliliter. BAU/mL = binding antibody units per milliliter.

